# Comprehending Atopic Risk Elements (CARE): An observational study to determine lifestyle, biophysical and environmental risk factors in the development of early-onset pediatric atopic dermatitis

**DOI:** 10.1101/2023.11.01.23297922

**Authors:** Michael Brandwein, Keren Gamrasni, Tamar Landau, Alex Levin, Tatiana Smolkin, Sofia Bauer-Rusek

## Abstract

**Background:** Atopic dermatitis and food allergies affect a growing swath of the population and there is consensus that their development is determined by a confluence of inherent and environmental factors. Of the numerous influences identified, a significant proportion of them are readily accessible from birth, thereby potentially opening a path for risk stratification from birth. The CARE study aims to harness this knowledge, coupled with advances in machine learning predictive modeling, to effectively determine whether a neonate is at-risk for developing atopic dermatitis or food allergies from birth.

**Methods & Design:** The CARE study is a prospective observational study of neonates recruited 1-5 days following birth from the neonatal ward of participating medical centers. Upon recruitment, trans-epidermal water loss measurements will be taken from neonates and their biological parents, and a survey will be administered to parents to record various environmental, historic and lifestyle elements that may contribute to or protect against the development of atopic dermatitis and food allergy. Follow-up questionnaires will be administered at ages 6, 12 and 24 months. Atopic dermatitis outcome measures, primarily a modified version of the UK Working Party diagnostic criteria for atopic eczema, will be assessed at 6, 12 and 24 months and food allergy outcome measures will be assessed at 12 and 24 months of age.

**Discussion:** The data generated from the CARE trial will serve to validate the notion that easily-accessible measures of risk can enable risk stratification from birth for infants at-risk of developing atopic dermatitis and food allergies.

**Trial Registration:** www.clinicaltrial.gov NCT04325451, prospectively registered on March 27, 2020

## Background

Atopic dermatitis (AD) is a chronic, relapsing inflammatory skin disorder which primarily affects children (1,2). The disease is now two to three times more prevalent in children than it was just four decades ago, yet the cause for its rise is not entirely clear (3). It is manifested by eczematous skin lesions associated with severe itch, leading to a significant impairment in quality of life. Children affected with AD report lower sleep quality (4), as do their mothers (5), in addition to a general decrease in quality of life (6). Treatment approaches include an array of topical and systemic agents, including hydrocortisones, antihistamines, emollients, anti-pruritics, topical corticosteroids, UV light therapy, immunomodulators, anti-inflammatory agents and monoclonal antibodies (3). While the current guidelines for the management of the condition recommends trigger avoidance, there is currently no modality to prevent disease incidence and/or occurrence.

The term “atopic march” has been coined to recognize the increased comorbidity of atopic dermatitis, allergic rhinitis and asthma, or the atopic triad. Additionally, although not an atopic condition, food allergy occurrence may be associated with the aforementioned conditions as well. The increased co-occurrence of these conditions has been shown in several studies and is independent of the association of the conditions with IgE sensitization (7,8). Due to the typical age of onset of AD (first two years of life), it has been posited that AD disease occurrence plays a pivotal role in the progression of the atopic march (2). Important strides have been taken in the last decade identifying early introduction of allergenic foods as an important food allergy risk mitigation strategy (9,10). These studies have been translated into practice through updated guidelines, and their particular relevance to at-risk populations is evident through both study results and the updated guidelines (11–13).

Skin barrier impairment has been identified as a leading risk factor for the development of AD. Filaggrin loss-of-function mutations increase the risk of contracting AD by more than threefold. Filaggrin is a filament-binding protein in the skin and its absence causes a defective skin barrier (14,15). Trans-epidermal water loss (TEWL) measurement is a non-invasive method to assess the integrity of the skin barrier (Figure 1). Elevated TEWL was demonstrated in AD patients on both affected and non-affected skin and its levels have been correlated with *Staphylococcus aureus*, a microbial pathogen long implicated in the pathogenesis of AD (16). High TEWL forehead measurements at birth was associated with AD development, and may be an important factor contributing towards the potential efficacy of preventative prophylactic emollient application on neonates (17).

**Figure 1:**
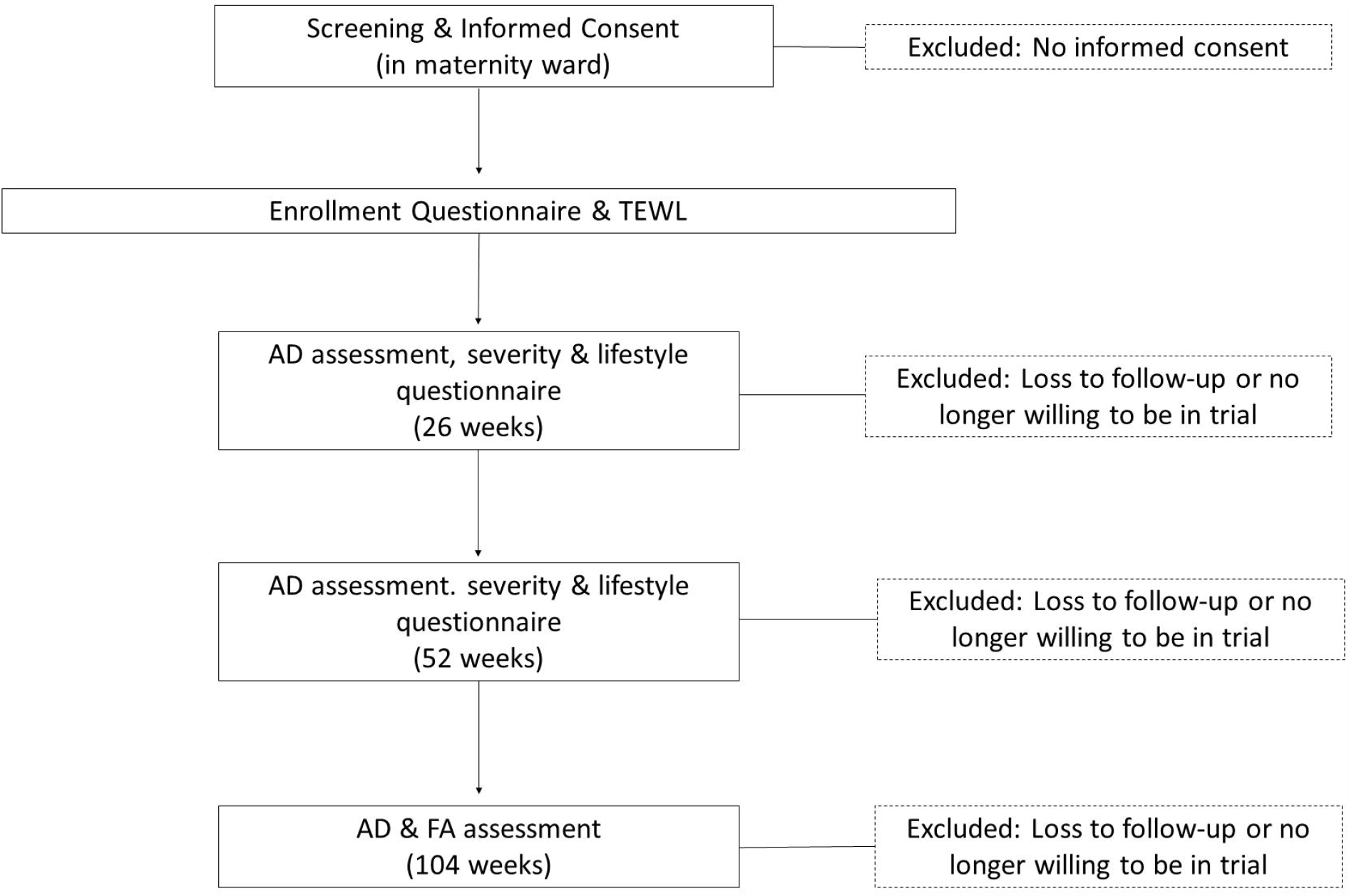
Skin Barrier Impairment & Trans-epidermal Water Loss: Skin barrier impairment is represented by a disruption in the orderly brick and mortar presentation of the epidermal cells (left panel). This subsequently leads to a larger loss of water from the body, measured through TEWL. In contrast, a tight skin barrier keeps water inside the body, thereby leading to a lower TEWL (right panel).

Additional lifestyle and environmental risk elements of both atopic dermatitis and food allergies have been documented extensively in the past few decades. Aside from familial history of atopy, other risk factors associated with a higher prevalence of these conditions include urban dwelling (18,19), household smoking (20,21), gestational age at birth, presence of household pets, birthweight, exposure to tobacco smoke, maternal age at birth, season of birth, frequency of contact with farm animals and others. However, despite the extensive body of research documenting these risk factors, they have yet to be synthesized into a tool that can stratify risk for AD, and therefore risk stratification in the clinical setting still relies primarily on familial history of atopy.

The goal of this trial is to recruit a birth cohort from the general population and assess their familial, lifestyle, environmental and skin barrier risk factors at birth continuing through two years of age. Atopic dermatitis and food allergy endpoint data will be collected as well. Participants’ data will then be used to validate an artificial intelligence empowered risk stratification algorithm described elsewhere.

## Methods & Design

### Aims

The primary aim of this study is to determine whether the combined information of familial history of atopy, TEWL, and other risk factors can effectively predict the risk of developing AD until two years of age. This will be accomplished by assessing the accuracy of a regression model, trained on an independent dataset, in stratifying risk in this prospective cohort. The secondary aim of the study is to determine whether the combined information of familial history of atopy, TEWL, and other risk factors can effectively predict the risk of developing FA at one year and two years of age.

### Design and setting

This is a multicenter, prospective observational longitudinal study. Participating medical centers include Meir Medical Center (Kfar Saba, Israel) and Poriya Medical Center (Tiberias, Israel). The patient population in these two medical centers represent a large swath of the population in Israel, with widely varying socio-economic levels and representation from both urban and rural communities. Neonates will be recruited from maternity wards amongst the general population within the first one to five days of life.

### Recruitment of study subjects

Mothers of neonates will be invited to enroll in the study by study coordinators during their postnatal stay at the hospital. The details of the protocol will be discussed with each neonate’s parent or legal guardian and written informed consent will be obtained for all subjects before any study-related procedure is performed. In obtaining informed consent, the information will be provided in language and terms understandable to the subject’s parents. The signed and dated consent form itself will be retained by the investigator as part of the trial records. A copy of the signed and dated consent form will be given to the subject’s parents or legal guardian, as appropriate.

### Study population

Term and near-term (34-42 weeks gestational age) healthy neonates delivered in a hospital setting between 24 and 120 hours prior to enrollment will be recruited. Mothers must be at least 18 years of age and the neonate’s parents must agree to complete the study questionnaires at the defined times throughout the study. Exclusion criteria for the study include birth before 34 weeks gestational age, neonate treated in an incubator in the 12 hours prior to study recruitment or having undergone or planning to undergo major surgery in the coming 14 days. Additionally, neonates with an existing widespread skin condition that would make the detection and/or assessment of eczema difficult, with a serious health issue which, at parent or investigator discretion, would make it difficult for the neonate or parents to take part in the trial, or who has been administered oral or parenteral antibiotics from birth until study enrollment will be excluded from the study.

### Data collection

TEWL measurements from the forearm and forehead will be taken from neonates and their biological parents at baseline. Additional lifestyle, environmental and historic data will be collected at baseline, and then again at 6, 12, and 24 months of age. These data include information on familial history of atopic conditions, demographics, household, medical history of the infant, duration of the pregnancy and feeding practices. A timeline of assessments is presented in Table 1 and the SPIRIT flow diagram is presented in Figure 2. The REDCap (Research Electronic data Capture) and OpenClinica data management platforms will be used for data collection and management (22,23).

**Table 1.**
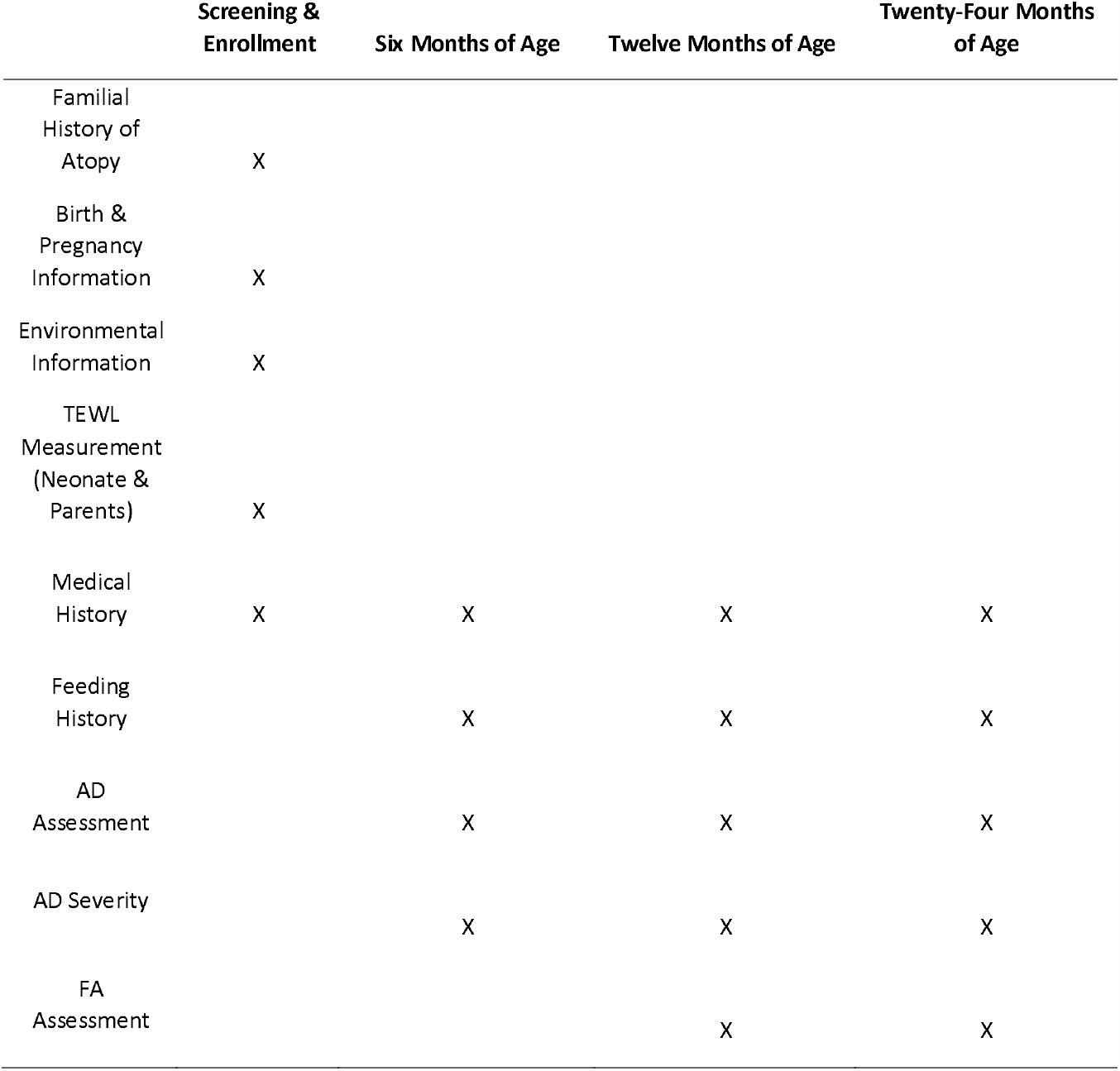
Summary of Assessments.

|  | Screening &<br>Enrollment | Six Months of Age | Twelve Months of Age | Twenty-Four Months<br>of Age |
| --- | --- | --- | --- | --- |
| Familial<br>History of<br>Atopy | X |  |  |  |
| Birth &<br>Pregnancy<br>Information | X |  |  |  |
| Environmental<br>Information | X |  |  |  |
| TEWL<br>Measurement<br>(Neonate &<br>Parents) | X |  |  |  |
| Medical<br>History | X | X | X | X |
| Feeding<br>History |  | X | X | X |
| AD<br>Assessment |  | X | X | X |
| AD Severity |  | X | X | X |
| FA<br>Assessment |  |  | X | X |

**Figure 2:**
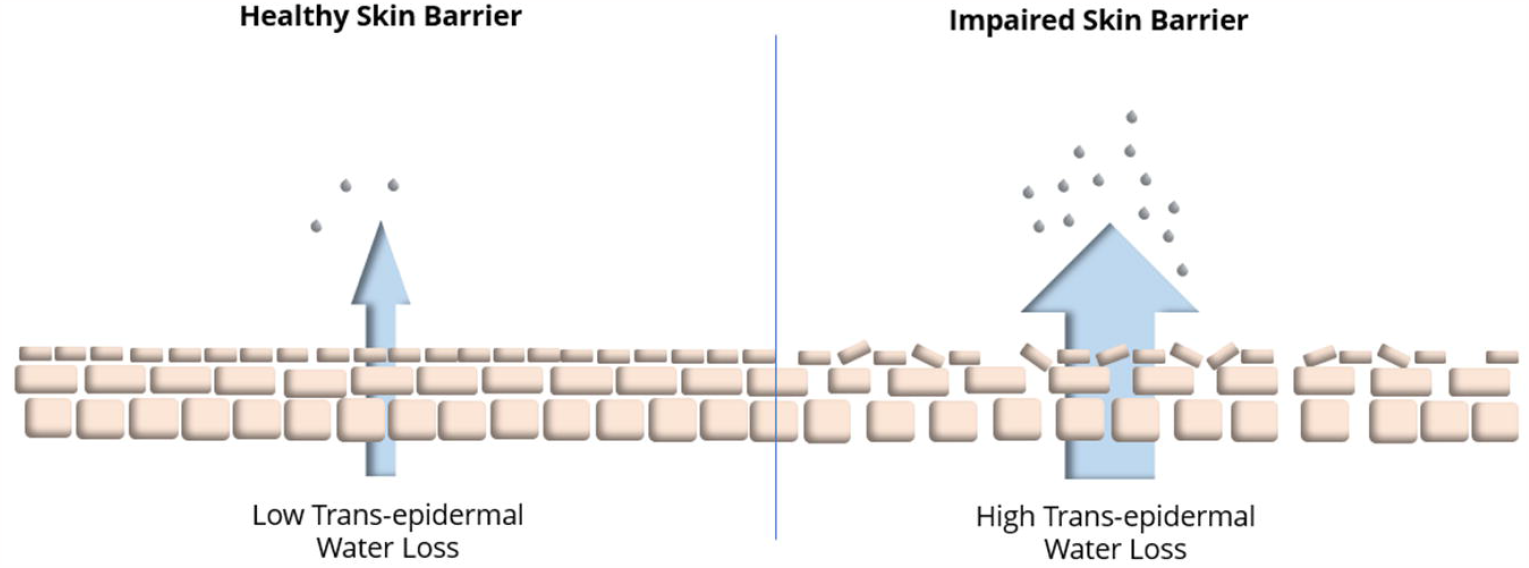
SPIRIT Flow Diagram. depicted study flow and procedures.

### Questionnaires and surveys

All parents of participants will be asked to fill out a questionnaire at study enrollment. The questionnaire documents the infant’s medical history, his or her familial setting and familial history of atopy, additional medical history pertaining to the mother and the pregnancy and the infant’s environment. An additional lifestyle questionnaire will be administered to parents of participants at 26 & 52 weeks from study enrollment. The interim questionnaire surveys medical history since the previous contact, infant feeding practices and additional environmental factors.

Cumulative incidence of AD will be evaluated using a modified version of the UK Working Party diagnostic criteria for atopic eczema (24). Evaluations will take place at 26 weeks, 52 weeks and 104 weeks from birth. In addition, cumulative incidence of AD will be assessed by parental report of a medical diagnosis by the infant’s pediatrician and/or dermatologist. Age of atopic dermatitis onset will be evaluated using the text of the validated questionnaire for disease onset of the UK Working Party diagnostic criteria for atopic eczema, with changes to the answers of question 2 to account for the age of the study population. AD severity will be evaluated using the Patient Oriented Eczema Measure (POEM) questionnaire and accompanying scoring criteria (25). Additionally, severity will be assessed by parental report of first steroidal indication by physician as part of the interim questionnaire at 26, 52 and 104 weeks from birth.

FA assessment will be evaluated based on the 2010 U.S. Food and Drug Administration Food Safety Survey Questionnaire, modified to reflect the age of infants in the study (26), at 52 and 104 weeks from study enrollment. Additionally, it will be assessed by parental report of pediatrician or allergist diagnosis.

### TEWL Measurement

After consent, baseline trans-epidermal water loss (TEWL) will be measured from the volar forearm and forehead in triplicate from each site using a Delfin Vapometer, a CE Class-II medical device. TEWL measurements will be taken from parents and patients alike. Patients who have been in the incubator will only be measured if they have been out of the incubator for at least 12 hours.

### Ethics and dissemination

This study will be conducted in accordance with the International Council for Harmonisation of Technical Requirements for Pharmaceuticals for Human Use guidelines (Assembly Helsinki, 1964 and all subsequent amendments)(27), and local Ministry of Health guidelines.

The study was approved by the Helsinki Committee of the Meir Medical Center, Kfar Sabba, Israel (approval number 241-20) and by the Helsinki Committee of the Poriya Medical Center, Tiberias, Israel (approval number 0107-19). Written informed consent is obtained by parents/guardians of newborns according to the approved protocols and procedures received from the respective Helsinki Committees. The study was registered on www.clinicaltrials.gov (Identifier: NCT04325451) on March 27, 2020, prior to subject recruitment.

Participants will be assigned a unique identifier by the investigator. The investigator will be requested to complete a linkage sheet that links each Patient Study Number with patient identifying information, in order to be able to retrieve the patient medical records in case of queries at the data quality control stage. The linkage sheet will be kept solely by the investigators and will not be shared with any third party.

No adverse events are anticipated as a result of the study as this is an observational study.

### Statistical methods

All measured variables and derived parameters will be listed individually and, if appropriate, tabulated by descriptive statistics. For categorical variables summary tables will be provided giving sample size, absolute and relative frequency and 95% CI (Confidence Interval) for proportions by study group. For continuous variables summary tables will be provided giving sample size, arithmetic mean, standard deviation, coefficient of variation (CV%), median, minimum and maximum and 95% CI (Confidence Interval) for means of variables by study group. All tests will be two-tailed, and a p-value of 5% or less will be considered statistically significant. The data will be analyzed using R. All supportive analyses and analyses of secondary/long term outcomes will be documented in the Statistical Analysis Plan which will be finalized prior to database lock. This will include methods to deal with missing data and sub-group analyses.

The primary aim of the study is to assess a previously developed machine-learning enabled algorithm’s accuracy in predicting the risk of developing atopic dermatitis. Multivariate Logistic Regression models will be applied for testing the statistical significance of groups of risk/protective factors in the cumulative incidence of AD at one year of age. Additionally, the Cox model will be applied for analysis of detecting difference in time to disease occurrence, with adjustment to possible covariates and the Kaplan-Meier survival function curve will be applied for testing the statistical significance of the difference in time to disease occurrence based on different risk and/or predictive factors.

## Discussion

This study will allow the prospective validation of an artificial intelligence empowered risk stratification algorithm for use on a neonate in the hospital or primary care setting. Additionally, it will examine whether the integration of birth TEWL measurements alongside well established risk factors provides additional predictive value to such a risk-stratification approach. Effective and early risk stratification can both inform the standard of care and potentially enhance the efficacy of prevention strategies by targeting those at-risk and thereby increasing compliance.

## Data Availability

Not applicable

## Abbreviations

AD: Atopic Dermatitis
CARE: Comprehending Atopic Risk Elements
FA: Food allergy
POEM: Patient Oriented Eczema Measurement
TEWL: Trans-epidermal water loss

## Declarations

### Consent to publish

Not applicable

### Availability of data and materials

Not applicable

### Authors’ contributions

MB wrote the manuscript and designed the study. KG & AL coordinated the implementation, data collection and execution of the study. TL contributed to the data preparation and analysis. TS & SB serve as co-principal investigators, contributed towards study design and recruit maternal-neonate dyads. All authors read, revised and approved the final version of this manuscript.

## Acknowledgements

The authors would like to thank the parents and neonates who are participating in this study. In addition, we acknowledge Nardeen Hassan, Tamra Nissim, Yaara Hoffman and Avital Diamant for assisting in ethics approval and communication with the ethics committees. We would like to additionally acknowledge Inbal Kessler for the preparation of figures for this manuscript.

## Author details

^1^MYOR Diagnostics ltd., Barket 29 Zichron Yaakov, Israel, ^2^Department of Neonatal Care, Baruch Padeh Poriya Medical Center, Lower Galilee, Israel, ^3^Department of Neonatal Care, Meir Medical Center, Clalit Health Services, Kfar Saba, Israel

